# Butyrate and related epigenetic changes link Parkinson’s disease to inflammatory bowel disease and depressive symptoms

**DOI:** 10.1101/2021.09.17.21263343

**Authors:** Aoji Xie, Elizabeth Ensink, Peipei Li, Juozas Gordevičius, Lee L. Marshall, Sonia George, J. Andrew Pospisilik, Velma T. E. Aho, Madelyn C. Houser, Pedro A. B. Pereira, Knut Rudi, Lars Paulin, Malú G. Tansey, Petri Auvinen, Patrik Brundin, Lena Brundin, Viviane Labrie, Filip Scheperjans

## Abstract

**Background:** The gut microbiome and its metabolites can impact brain health and are altered in Parkinson’s disease (PD) patients. It has been recently demonstrated that PD patients have reduced fecal levels of the potent epigenetic modulator butyrate and its bacterial producers. Here, we investigate whether the changes in the gut microbiome and associated metabolites are linked to PD symptoms and epigenetic markers in leucocytes and neurons.

**Methods:** Stool, whole blood samples, and clinical data were collected from 55 PD patients and 55 controls. We performed DNA methylation analysis on whole blood samples and analyzed the results in relation to fecal short-chain fatty acid concentrations and microbiota composition. In another cohort, prefrontal cortex neurons were isolated from control and PD brains. We identified the genome-wide DNA methylation by targeted bisulfite sequencing.

**Results:** We show that lower fecal butyrate and reduced *Roseburia, Romboutsia*, and *Prevotella* counts are linked to depressive symptoms in PD patients. Genes containing butyrate-associated methylation sites include PD risk genes and significantly overlap with sites epigenetically altered in PD blood leucocytes, predominantly neutrophils, and in brain neurons, relative to controls. Moreover, butyrate-associated methylated-DNA (mDNA) regions in PD overlap with those altered in gastrointestinal, autoimmune, and psychiatric diseases.

**Conclusions:** Decreased levels of bacterially produced butyrate are linked to epigenetic changes in leucocytes and neurons from PD patients, and to the severity of their depressive symptoms. PD shares common butyrate-dependent epigenetic changes with certain gastrointestinal and psychiatric disorders, which could be relevant for their epidemiological linkage.

## Background

Parkinson’s disease (PD) is a neurodegenerative disorder that is typically characterized by motor impairments due to the death of dopaminergic (DA) neurons located within the substantia nigra pars compacta (SNpc) [1]. A hallmark of PD is the aggregation of misfolded α-synuclein protein in both the central and peripheral nervous system, forming aggregates or Lewy bodies [2]. The discovery of α-synuclein aggregates in the enteric nervous system, coupled with the early gastrointestinal (GI) symptoms of PD (e.g. constipation), has led to the hypothesis that the pathogenesis of PD may originate in the GI tract, or at least outside of the central nervous system [3]. Additionally, it is hypothesized that α-synuclein aggregates can be transported in a prion-like manner from the enteric nervous system to the central nervous system through the vagus nerve [4]. This hypothesis is supported by the decreased risk for PD amongst patients who have undergone a vagotomy [5]. Further intertwining the GI system with PD is the finding that the presence of gut microbiome is required for mice that overexpress α-synuclein to develop motor deficits [6]. These same mice displayed increased motor deficits after receiving a fecal transplant from a PD patient compared to receiving transplant from a healthy donor [6]. Moreover, the composition and function of the gut microbiome in PD patients has been shown to be significantly different from healthy controls in multiple studies [7–9]. While research has consistently shown a difference in the PD microbiome compared to the healthy microbiome, there is no consensus on the exact differences or what effect this dysbiosis might have on the PD patient [9].

Among the common differences seen in the PD microbiome across multiple studies is a decrease in bacteria which produce short-chain fatty acids (SCFAs) [10]. SCFAs are saturated fatty acids produced via the fermentation of dietary fiber by certain colonic bacteria [11]. The deficiency of SCFAs has been implicated in multiple diseases such as autoimmune disorders, cancer, metabolic syndromes, and neurological disorders [12]. Fecal samples from PD patients have been shown to harbor significantly lower concentrations of the SCFAs acetate, propionate, and butyrate when compared to healthy controls [13]. Butyrate, specifically, has been implicated as an important bacterial metabolite due to its role as a strong endogenous histone deacetylase (HDAC) inhibitor indirectly affecting DNA-methylation [14], allowing it to epigenetically alter the gene expression of multiple cell types [15]. Several studies have demonstrated the ability of butyrate to reduce the inflammatory properties of both innate and adaptive immune cells through inhibiting reactive oxygen species release, inflammatory cytokine production, and inducing activated immune cell apoptotic mechanisms [16–18]. In addition to its immune modulating properties, butyrate has been shown in multiple in-vivo studies to influence the CNS through decreasing blood-brain barrier permeability, decreasing microglial activation, and relieving anxiety and depression, which are both common prodromal PD symptoms. Butyrate has also shown neuroprotective effects in PD mouse models [19, 20]. However, the possible impact of the low butyrate level observed in PD patients on epigenome status as well as on clinical symptoms has yet to be elucidated.

To test whether butyrate impacts epigenetic markers in the blood and brain of PD patients, and whether this is associated with symptom severity, we performed DNA methylation profiling in whole blood samples and neuronal tissue from two cohorts of PD patients and controls (Fig 1), and related the findings to fecal gut microbiome and metabolite data and clinical symptoms.

**Fig 1:**
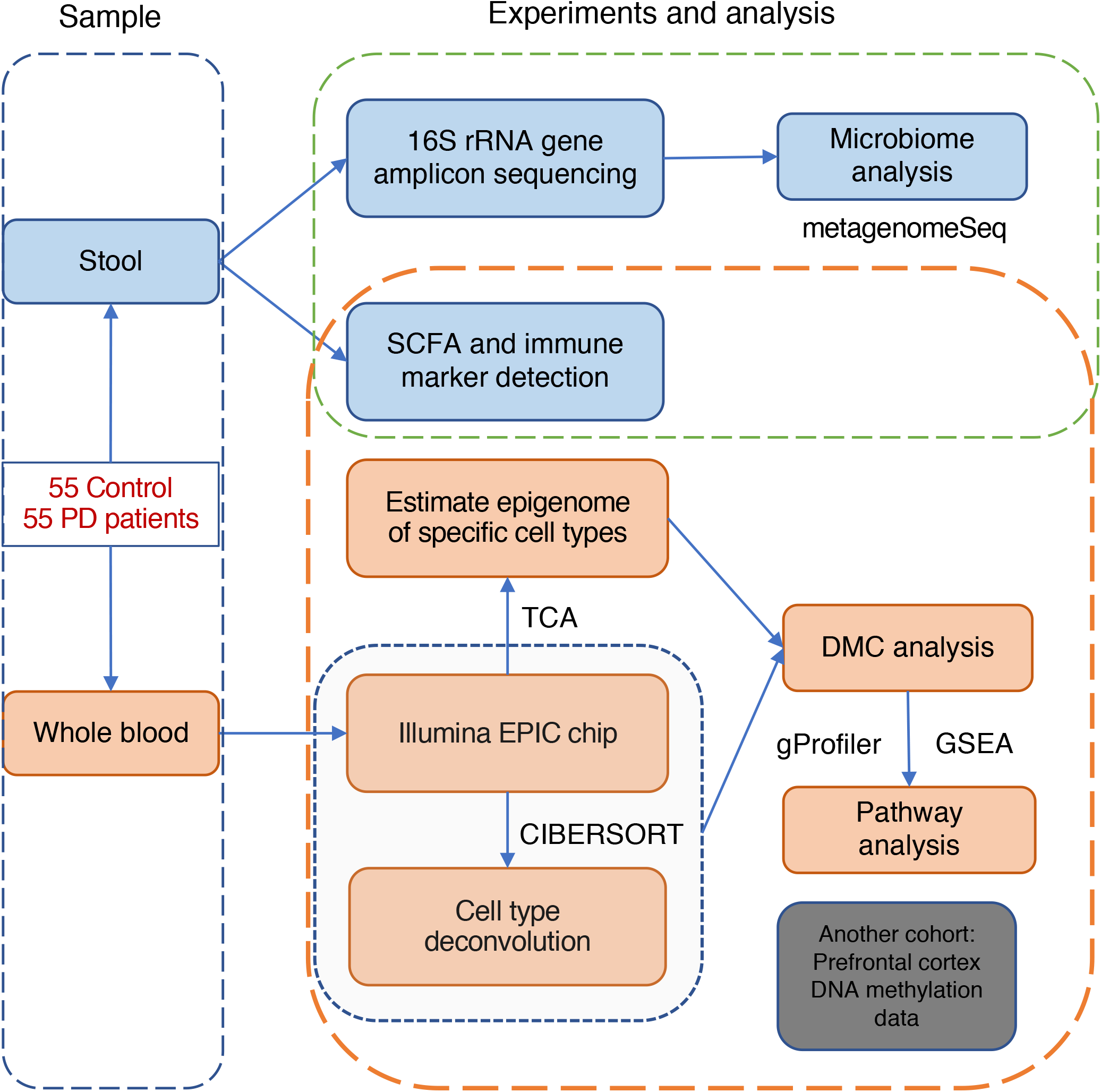
Outline of this study.

## Methods

### Human samples and metadata

Blood samples, clinical data, microbiome count data, inflammatory and permeability markers, and stool SCFA levels used in this study are from the Helsinki Parkinson microbiome cohort, and methodology of sampling, sample processing, and analysis have been described previously [21, 22]. The study was approved by the ethics committee of the Hospital District of Helsinki and Uusimaa. All participants gave informed consent. Human prefrontal cortex tissue for this study was obtained from the Parkinson’s UK Brain Bank, NIH NeuroBioBank, and Michigan Brain Bank, with approval from the ethics committee of the Van Andel Research Institute (IRB #15025). Methodology of sampling, sample processing, and analysis have been described previously [23]. We used the differential methylated cytosines data (supplementary file 5) in our analysis.

### Statistical Analyses

We performed statistical analyses with R [24] (v3.6.1) with packages including metagenomeseq [25] (v 1.27.3) for differential microbial data comparisons; ChAMP [26] (v 2.20.1), and minfi [27] (v 1.31.1) for EPIC array analysis; Tensor Composition Analysis (TCA) [28] (v 1.1.0) for epigenome estimation for immune cell types; limma [29] (v 3.41.17) for robust linear regression; ggplot2 [30] (v 3.3.1) for data visualization. All the code used for this work is publicly available: Github: https://github.com/AojiXie/PD_microbiome_DNA_methylation

### Genome-wide DNA methylation profiling

Whole-genome DNA methylation profiling for each sample was performed on Illumina Methylation EPIC BeadChip microarrays at Van Andel Institute Genomic core. Bisulfite converted DNA samples (n = 136, including replicates) were randomized across arrays (8 samples/array). Data generated from the microarrays were preprocessed with Minfi (v 1.31.1). Normalization was performed with noob [31]. We confirmed that the sex of the individuals matched that inferred from the DNA methylome (minfi getSex() function).

The portions of immune cell types (CD8^+^T cell, CD4^+^T cell, B cell, NK, monocyte, and neutrophil) were estimated by CIBERSORT [32] (Fig S2a) using whole blood-specific markers as reference [33]. The filtering method in ChAMP was used to filter probes: Probes that overlapped SNPs [34] (minor allele frequency > 0.05) on the CpG or single-base extension (95,485 probes), probes that aligned to multiple locations (42,558 probes), probes with a beadcount < 3 in at least 5% of samples (3380 probes), MultiHit Start [35] (11 probes), probes located on X,Y chromosome (16,541 probes), NoCG Start (2953), and those that failed detectability (p > 0.01) (7,951 probes) were excluded. After processing, 739,597 probes were left. Champ.svd() in ChAMP was used to test the batch effects. Those batch effects (array and slide positions) were corrected by ComBat[36]. After batch effects correction, the M value was ready for the subsequent statistical analysis. Cell-type-specific resolution epigenetics were performed using tensor composition analysis [28].

### Statistical analysis for differentially methylated sites

DNA methylation analysis involved robust linear regression models with empirical Bayes from the limma (v 3.41.17) statistical package [29]. P-values were adjusted with a Benjamini-Hochberg correction for multiple testing and those with FDR *q* < 0.05 were deemed significant.

### Model

#### 1: In whole blood methylation epigenome

Variable selection: Cell type percentages were used as covariates because the whole blood contains several immune cell types in varying proportions. This variation might affect the interpretations of DNA methylation levels based on whole blood DNA[37]. BMI and smoking history are used as covariates because their associations with whole blood DNA methylation have been observed [38, 39].

M value ∼ Butyrate + Age + Sex + Smoking history + BMI + CD4^+^T cell + CD8^+^T cell + B cell + Monocyte + Neutrophil

#### 2: In cell specific epigenome

M value ∼ Butyrate + Age + Sex + Smoking history + BMI

### Pathway enrichment analysis

To identify proximal interactions with gene targets, we used the GREAT (v4.0.4) software [40]. Gene annotation was performed for the gene targets of the significant cytosine sites in our analysis and for the background, consisting of gene targets for all cytosines included in our analysis. The background was 18,455 genes. Pathway analysis of methylated cytosines altered in PD and correlated with butyrate level was performed using g:Profiler [41], with networks determined by EnrichmentMap and clustered by AutoAnnotate in Cytoscape (v3.7.1) [42].

Since enhancer elements dynamically regulate gene expression through three-dimensional physical interactions, we analyzed chromatin interaction data to reveal the gene targets of enhancers relevant to the differential methylation sites affected by butyrate. For this analysis, we used promoter-centric chromatin interactions identified in blood cell types [43]. Gene enrichment set test of different blood cell types was performed by GESA [44, 45] (https://www.gsea-msigdb.org/gsea/msigdb/index.jsp) Reactome gene sets. Benjamini-Hochberg FDR *q* < 0.01 with minimal gene set less than 100 were used as the significant threshold.

### Genetic-epigenetic correlation

Genetic-epigenetic analysis was performed using LD Score (LDSC) software [46, 47], to estimate the correlations between butyrate-associated mDNA regions and the GWAS summary statistics of other diseases. To construct butyrate-associated mDNA regions for linkage disequilibrium (LD) score regression analysis, SNPs within +-5000bp of EPIC chip array sites were included, and the p-values of methylation cytosines in butyrate linear model were assigned to those SNPs. If a SNP was within +-5000bp of more than one methylated cytosine, the smallest p-value was selected. The summary statistic of a 2019 PD GWAS study [48] was used in this analysis. For other diseases, we used the summary statistics which are provided in the LD Hub interface [49]. P < 0.05 was used as the significance threshold.

The common SNPs (with p < 0.05) between butyrate-associated mDNA regions, PD GWAS summary statistics, and other diseases, respectively, were extracted. GREAT [40] was used to get the gene annotation of those common SNPs with Association rule: Basal + extension: 5000 bp upstream, 1000 bp downstream, 600000 bp max extension, curated regulatory domains included. Gene set enrichment analysis was performed by GSEA using Reactome gene sets [44, 45] (https://www.gsea-msigdb.org/gsea/msigdb/index.jsp). Benjamini-Hochberg FDR *q* < 0.01 with minimal gene set less than 100 were used as the significant threshold.

## Results

### Butyrate-producing microbes are altered in PD and correlate with depressive symptoms

First, we re-analyzed previously published [21, 22] raw data on SCFA levels and 16S rRNA gene amplicon counts from stool samples of PD patients and healthy controls. The metagenomeSeq was used for differential microbial analysis [25]. Unlike RNAseq studies, most OTUs are rare (absent from a large number of samples) because of insufficient sequencing depth (under sampling) or some organisms only being present in few samples. This sparsity can lead to strong biases when sequence read counts are tested for significant differences. Zero counts in samples with low coverage are misinterpreted as absent taxonomic features. The advantages of metagenomeSeq is the zero-inflated Gaussian mixture (ZIG) model which removes testing biases resulting from under sampling and the cumulative-sum scaling (CSS) normalization method to avoid biases of uneven sequencing depth [50]. We can confirm significantly reduced butyrate levels (Benjamini-Hochberg FDR *q* < 0.05, robust linear regression) (Fig S1a) and differential abundances of *Bifidobacterium, Butyricicoccus, Clostridium_XlVa, Lactobacillus, Prevotella*, and *Roseburia* in PD patients (Benjamini-Hochberg FDR *q* < 0.05, metagenomeSeq ZIG model) (Fig S1b) [21]. Important new findings within the PD group are links between depressive symptoms as measured using the Geriatric Depression Scale (GDS-15) and lower fecal butyrate levels (Benjamini-Hochberg FDR *q* < 0.05, robust linear regression) (Fig 2a) as well as lower counts of *Prevotella, Romboutsia*, and *Roseburia* and higher counts of *Deltaproteobacteria_unclassified* (Benjamini-Hochberg FDR *q* < 0.05, metagenomeSeq ZIG model) (Fig. S1c, 2b). In line with these findings, we confirm a positive correlation of *Romboutsia* and *Roseburia* with butyrate levels in PD patients (Fig.S1c) [22]. In addition, no other symptoms including gastrointestinal (Rome-III questionnaire; Wexner score), motor symptoms (the Unified Parkinson’s Disease Rating Scale, UPDRS), nor non-motor symptoms (Non-Motor Symptoms Questionnaire, NMSQ; Non-Motor Symptoms Scale, NMSS) were linked with butyrate in PD patients.

**Fig 2:**
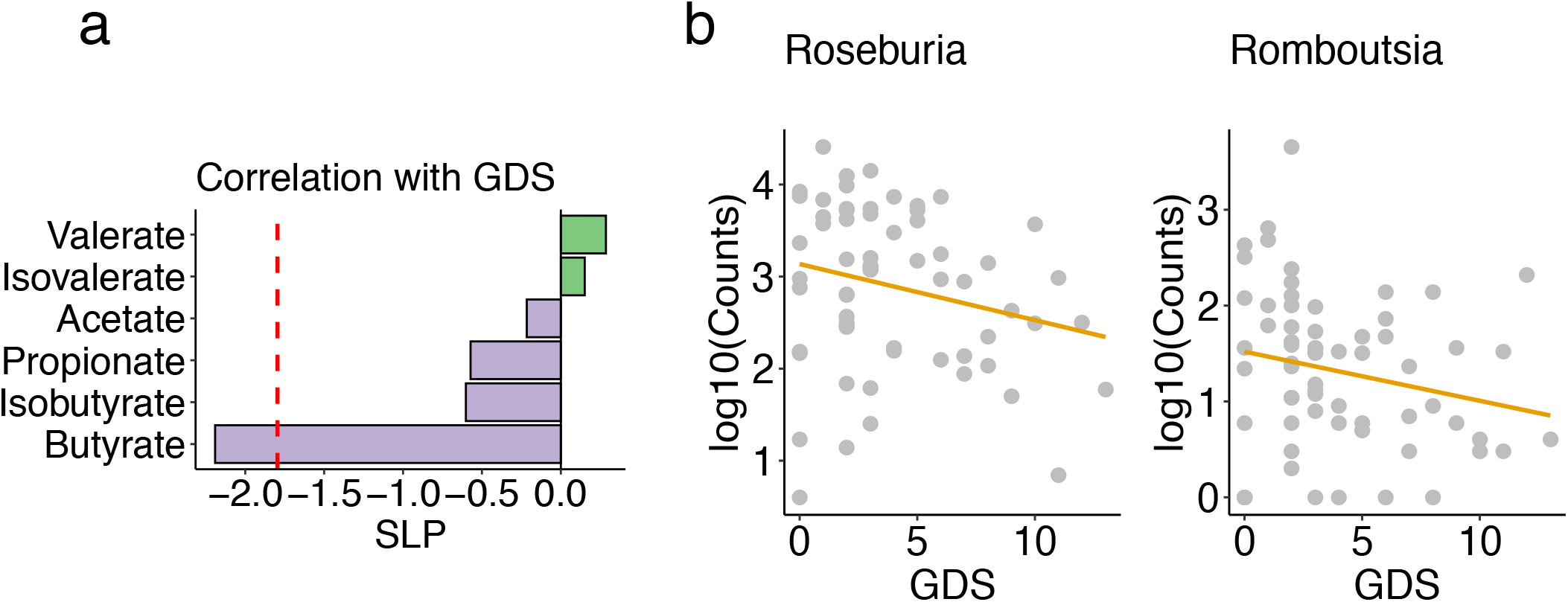
Butyrate and butyrate-producing microbes are associated with PD depressive symptoms. A: Short-chain fatty acids change with Geriatric Depression Scale (GDS) total scores in PD patients (robust linear regression, adjusting for age, sex, smoking status and BMI. Benjamini-Hochberg FDR *q* < 0.05 is used as the significant threshold). B: The butyrate-producing bacterial genera *Roseburia* and *Romboutsia* were negatively associated with GDS total scores in PD patients. Benjamini-Hochberg FDR *q* < 0.05, signed logP: SLP.

### Fecal butyrate levels are associated with epigenetic alterations in leucocytes and neurons of PD patients

We identified 3195 CpG sites that correlated significantly with stool butyrate levels in PD patients (Benjamini-Hochberg FDR *q* < 0.05, robust linear regression) (Fig. 3a) and 2950 CpG sites that were significantly changed in PD relative to control (Benjamini-Hochberg FDR *q* < 0.05, robust linear regression). In a previous study, we identified genes epigenetically altered in cortical neurons from PD patients [23]. Genes containing the butyrate-associated methylated cytosines in blood cells significantly overlapped with those genes altered in PD patients’ blood cells (relative to controls) and with those genes altered in PD patients’ prefrontal cortex neurons (relative to controls) (genes containing modified cytosines at Benjamini-Hochberg FDR *q* < 0.05, Fisher’s exact test *p* < 0.05) (Fig. 3b, c). Pathways involving those genes epigenetically altered in PD or with butyrate were identified and included neurodevelopment, cell development, synaptic transmission, metabolism, and signal transduction (Fig. 3d).

**Fig 3:**
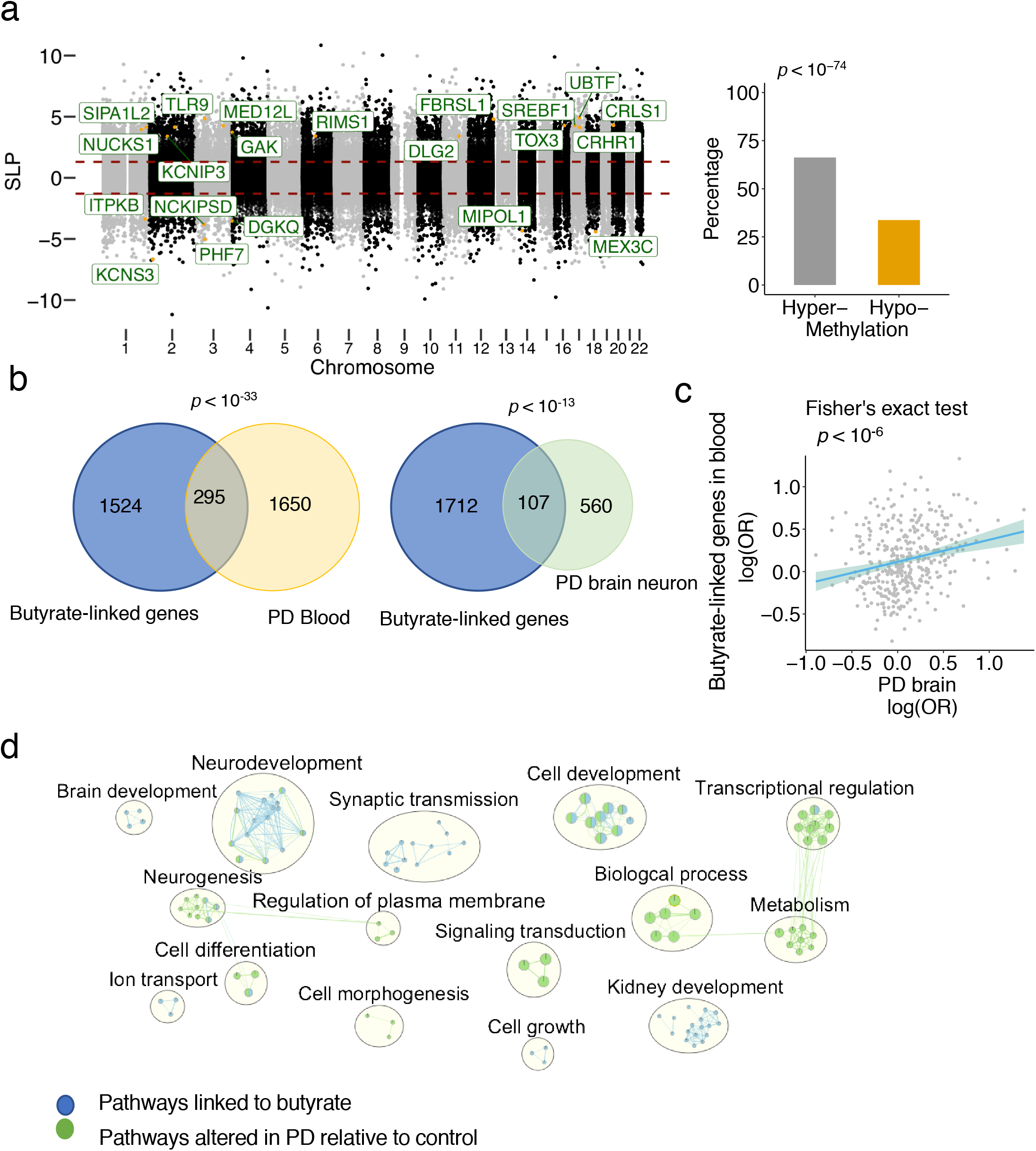
Links of butyrate to PD epigenome in brain and blood. DNA methylation analysis was performed by Illumina EPIC array in PD and control whole blood (n = 55 PD, 55 controls). a: DNA methylation changes in the blood of PD patients associated with butyrate levels are identified (robust linear regression, 3195 cytosines at Benjamini-Hochberg FDR *q* < 0.05, after adjusting for age, sex, smoking status, BMI and blood cell types). PD risk genes (identified by GWAS) with differential methylation are highlighted. The percentage of hyper-methylated and hypo-methylated cytosines is plotted (Fisher’s test, *p* < 0.05), butyrate level is positively associated with cytosine methylation in PD. b: Genes in PD blood (295 genes) and PD prefrontal cortex (107 genes) are significantly converged on the genes linked to butyrate levels (Fisher’s exact test, *p* < 0.05) c: Epigenetic alterations in blood and brain converge on those methylation sites linked to butyrate. The log (odds ratio) of genes in PD blood is significantly correlated with the log(odds ratio) of those genes in PD prefrontal cortex (Fisher’s exact test, *p* < 0.05). d: Enrichment analysis of genes epigenetically linked to butyrate and altered in PD relative to control. Nodes are *q* < 0.05 pathways merged by EnrichmentMap in Cytoscape.

### Epigenetic links between butyrate and leucocytes differ between cell types

To test if leucocyte epigenetic changes are associated with fecal butyrate, we used cell type-specific resolution epigenetics, TCA [28], to analyze the epigenome for each cell type (Fig S2). To test the epigenetic links of butyrate on different immune cell types, we analyzed the significant methylation sites altered by butyrate in the epigenome of neutrophils, monocytes, CD8^+^ T cells, CD4^+^ T cells, and B cells. All cell types were linked differently to butyrate, with neutrophils and monocytes containing the largest number of genes that were epigenetically linked to butyrate (Fig. 4a). Levels of inflammatory stool cytokines were derived from a previously published study of the same samples [22]. We also analyzed the methylated cytosines that were correlated with blood inflammatory cytokine levels. There was a large overlap between the genes epigenetically altered by butyrate and the genes epigenetically associated with levels of inflammatory cytokines (TNF, IL6, CXCL8, IL4, IL1B, IL10, IFNg, IL13, IL12p70, IL2 and LBP) in both monocytes and neutrophils (Fisher’s exact test, *p* < 0.05) (Fig. 4b). Pathway analysis shows that epigenetic alterations of three recently identified PD polygenic risk pathways [51] (neutrophil degranulation, metabolism of lipids, and innate immune system) in the monocyte and neutrophil epigenome were linked to butyrate levels (Fig. 4c). We also analyzed the effects of other SCFAs, and our results showed that the genes epigenetically altered in neutrophils were most strongly associated with butyrate compared to other SCFAs (Fig. S3a) and that there were no significant PD polygenic risk pathways altered by other SCFAs in neutrophils (Fig. S3b). These results indicate that neutrophil and monocyte functions are specifically altered in PD patients and that they might be epigenetically altered by butyrate.

**Fig 4:**
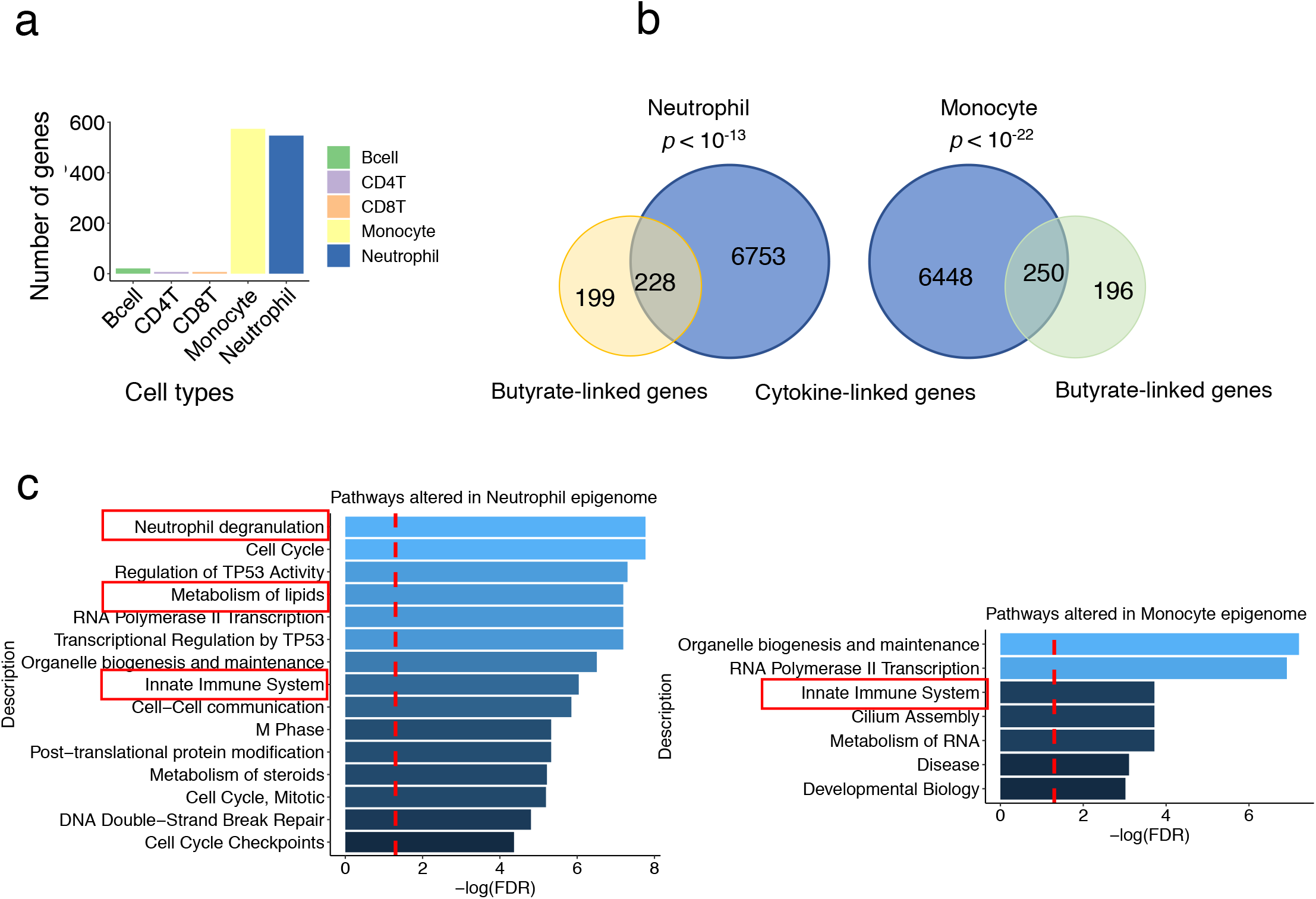
Blood cell types are differentially affected by butyrate. a: Significant methylation sites (related gene numbers) altered by butyrate in the epigenome of neutrophils, monocytes, CD8^+^T cells, CD4^+^T cells, and B cells, respectively. b: epigenetically altered genes linked to cytokines overlap with those epigenetically linked to butyrate in monocytes and neutrophils. Fisher’s exact test *p* < 0.05. c: Gene set enrichment analysis on the monocyte and neutrophil genes that are epigenetically altered and linked to butyrate. PD polygenic risk pathways including innate immune systems, metabolism of lipids and neutrophil degranulation are significantly altered. Significant threshold Benjamini-Hochberg FDR *q* < 0.05.

### Epigenetic-genetic correlation between diseases and butyrate-associated methylated-DNA (mDNA) regions

To investigate the epigenetic-genetic correlation, we ran a linear regression model to compare the results of methylation analysis to the GWAS of the odds-risk ratios for PD and other diseases, including gastrointestinal (ulcerative colitis, Crohn’s disease), autoimmune (rheumatoid arthritis), neurodegenerative (Alzheimer’s disease), and psychiatric diseases (bipolar disorder, schizophrenia, and major depression) and our DNA methylation study, adjusting for LD scores. We found that butyrate-associated mDNA regions are most strongly related to GWAS loci linked to PD and inflammatory bowel diseases. Further significant links were found with the GWAS loci of rheumatoid arthritis and bipolar disorder (linear regression, *p* < 0.05) (Fig. 5a). In contrast, Alzheimer’s disease, schizophrenia, or major depression GWAS loci were not linked to butyrate-associated mDNA regions. Pathway analysis was performed on the common genomic regions (butyrate-associated mDNA regions, GWAS loci of PD and each of the other diseases, respectively). All of the six PD polygenic risk pathways without known PD risk loci [51] were significantly altered in ulcerative colitis, Crohn’s disease, rheumatoid arthritis, and bipolar disorder (FDR *q* < 0.05) (Fig. 5b). The top 15 significant pathways are shown in Fig. S4. These results indicate that butyrate-associated epigenetic changes may contribute to observed epidemiologic links between PD and gastrointestinal, autoimmune, as well as certain psychiatric diseases.

**Fig 5:**
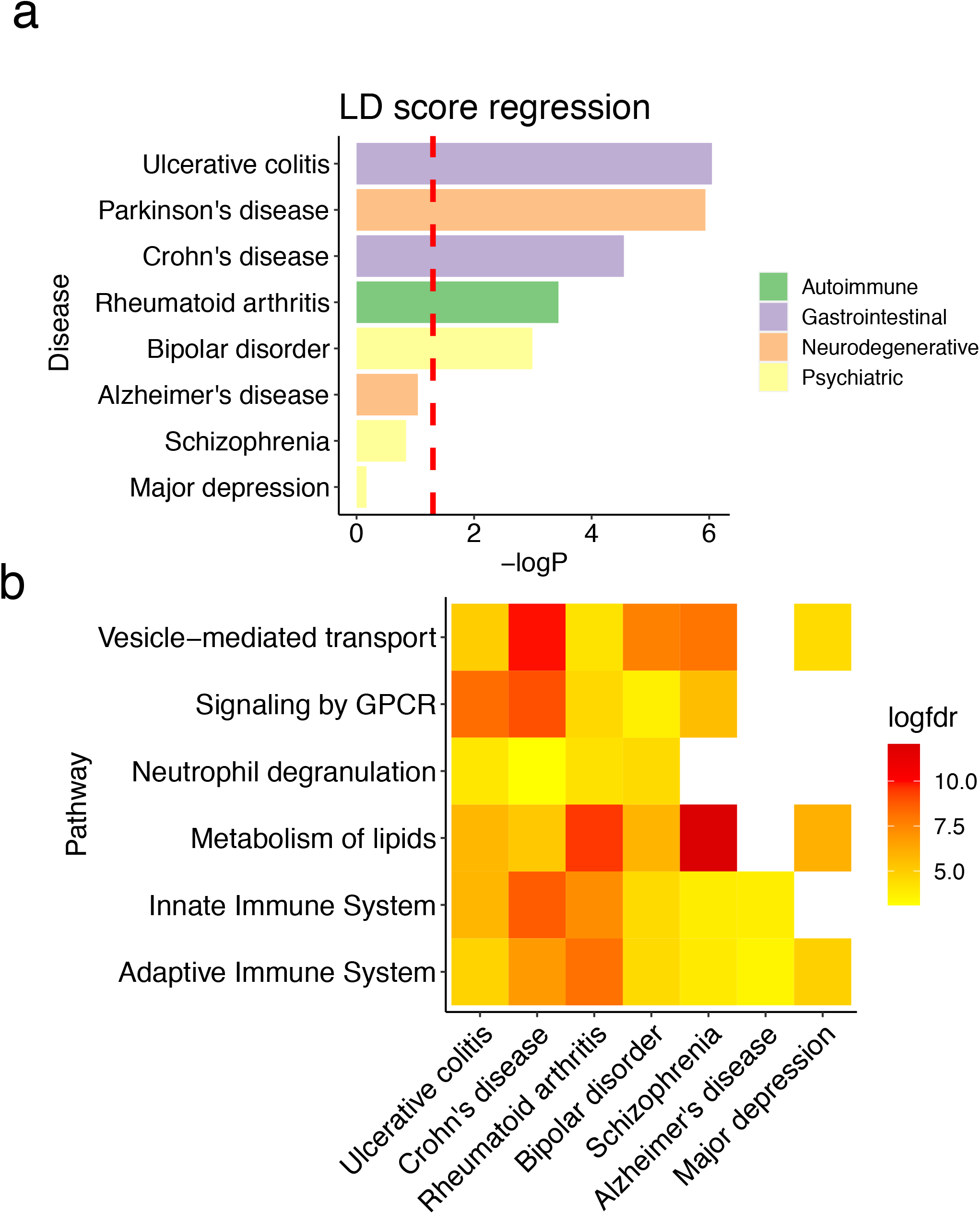
Epigenetic-genetic correlation between butyrate associated mDNA regions and the GWAS of other diseases. a: LD score regression was performed between butyrate associated mDNA regions and the GWAS summary statistics of Parkinson’s disease, ulcerative colitis, Crohn’s disease, rheumatoid arthritis, bipolar disorder, Alzheimer’s disease, schizophrenia and major depression, respectively. *p* < 0.05 is used as the significant threshold. b: Gene set enrichment analysis on the location of common genetic regions in both Parkinson’s disease, butyrate associated mDNA regions and those diseases in panel a. The log(FDR) of 6 PD polygenetic risk pathways without known PD risk loci [41]were plotted. Benjamini-Hochberg FDR *q* < 0.05 are considered as significant threshold, white squares indicate non-significant associations.

## Discussion

Several studies have found alterations of microbial composition and metabolites in PD, but the mechanisms linking these changes to PD and its symptoms are not well understood [9]. The bacterial metabolite butyrate is a strong HDAC inhibitor, with known epigenetic effects [52]. Several studies have shown the ability of butyrate to affect the innate and adaptive immune system as well as to alter the blood-brain barrier permeability [19, 53]. This study is the first to examine the possible role of epigenetic changes as a link between gut microbiota, its metabolites, and the pathophysiology of neural and immune cells in PD.

In this study, genome-wide DNA methylation profiling revealed significant epigenetic changes in the leucocytes and neurons of PD patients that overlap with genes the methylation of which is linked to fecal butyrate levels, including loci in PD risk genes [23]. Furthermore, butyrate levels correlated most strongly with abundance of bacteria belonging to the genera *Romboutsia* and *Roseburia*, known SCFA producers.

A major finding of our study is that the epigenome of specific immune cell types in PD is differentially linked to fecal butyrate levels. In particular, neutrophil and monocyte epigenomes show the strongest links to butyrate. We have also shown that the neutrophil epigenome is linked less to other SCFAs. A recent study [51] reported that neutrophil degranulation is potentially linked to PD risk. Our gene set enrichment analysis suggests that butyrate-linked epigenetic changes may impact this pathway. Several studies have reported an increase of neutrophils in PD patients, including studies using flow cytometric and epigenetic profiling approaches [54], which is consistent with our cell deconvolution results from the methylation profiling. However, our study for the first time decomposed the estimated epigenome of each cell type and investigated the epigenetic change from different immune cell types in PD and in connection to SCFAs. Our results suggest that butyrate may impact PD through epigenetic effects on innate immune cells and PD-related genes.

The exact dynamics that link fecal butyrate levels with blood leucocyte and brain epigenetic are not known. While a significant proportion of microbial-released butyrate is rapidly taken up and consumed locally in the gut, butyrate can cross the epithelial barrier and enter the circulation via the hepatic portal vein [55]. Microbiota-derived butyrate impacts histone acetylation in multiple tissues [56]. While concentrations in the portal vein are still considerable, concentrations in peripheral blood appear to be relatively low [55]. Thus, leucocytes get exposed to butyrate mostly in the gut wall and portal vein, while the impact of butyrate in systemic venous blood can be expected to be less and influenced by liver function. Epigenetic changes in blood leucocytes may impact inflammation systemically and in the brain [19]. At physiological concentrations, butyrate’s impact on brain metabolism and hippocampal neurogenesis has been shown in pigs [57]. While our findings support the importance of epigenetic mechanisms, their relative impact on the physiological effects of butyrate in the brain as compared to other mechanisms remains to be established.

Interestingly, our results suggest that patterns of butyrate-related epigenetic changes in PD are most similar to those found in inflammatory bowel disease and clearly less similar to those found in Alzheimer’s disease. Bowel diseases, in particular inflammatory bowel diseases [58], have been linked to an increased risk of PD and Alzheimer’s disease, but associations are stronger for PD [59]. Our results suggest that microbiome-related epigenetic modulation could be a mechanism linking gastrointestinal disorders and PD. Also, bipolar disorder [60] has been linked to an increased PD risk, and our results support a role for common epigenetic mechanisms in this context. In contrast, we could not find significant overlap with epigenetic patterns found in schizophrenia [61] and depression [62] which points to a lesser impact of epigenetics linking these disorders to PD. Interestingly, some overlap was seen with epigenetic patterns of rheumatoid arthritis, which reportedly is associated with a decreased PD risk [63–65]. Rheumatoid arthritis has also been linked to the gut microbiome, but changes have been somewhat opposite to those seen in PD, e.g. increase of *Prevotella* abundance in arthritis but decrease in PD [66].

In this study, we partly reanalyzed microbiome and metabolite data with methods not used in the previous publications [21, 22]. We observed that fecal bacterial butyrate is inversely correlated with depressive symptoms (GDS-15) in PD patients. Although there is no correlation between bacterial butyrate and other non-motor symptoms-related scales which include depressive related items (UPDRS I, NMSS and NMSQ), GDS-15 is more specific for assessing depressive symptoms. While we were able to reproduce PD-related microbiota alterations and decreased butyrate levels identified using earlier methods, we gained important new insights. Fecal butyrate and counts of *Prevotella, Romboutsia*, and *Roseburia* were negatively correlated with depressive symptoms in PD patients, potentially implicating bacterial metabolites in this important non-motor PD symptom.

## Conclusions

In sum, combining metabolite, microbiome, clinical data, and DNA methylation profiling, our study for the first time reveals a possible link between gut microbiome metabolite production and epigenetic changes implicating immune and neural pathways in PD patients with potential impact on depressive symptoms. Furthermore, our results point to microbiota-dependent epigenetic modulation as a potential pathway linking inflammatory bowel diseases and PD. Further research of altered bacterial metabolism and its impact on host physiology may reveal new biomarkers and therapeutic targets for PD.

## Data Availability

Microbiota data are available at the European Nucleotide Archive (accession number PRJEB27564) (https://www.ebi.ac.uk/ena/browser/view/PRJEB27564). Other data and files utilized in this study are available from the corresponding authors upon reasonable request.

## Ethics approval and consent to participate

The study was conducted according to the guidelines of the Declaration of Helsinki and approved by the ethics committees of the Van Andel Research Institute and the Hospital District of Helsinki and Uusimaa. All participants gave written informed consent.

## Consent for publication

Not applicable

## Availability of data and materials

### List of abbreviations

PD: Parkinson’s disease
SCFAs: Short-chain fatty acids
BMI: Body mass index
TCA: Tensor composition analysis
LD: linkage disequilibrium
UPDRS: Unified Parkinson’s Disease Rating Scale
NMSQ: Non-Motor Symptoms Questionnaire
NMSS: Non-Motor Symptoms Scale
GDS-15: Geriatric Depression Scale
GSEA: Gene set enrichment analysis

## Authors’ contributions

Conceptualization, AX, EE, PL, JG, LLM, and VL; data curation, EE, VTEA, MCH, KR, LP, PABP, LP, MGT, PA and FS; formal analysis, AX; funding acquisition, JAP, PB, LB, FS and VL; investigation, PB, LB, FS and VL; methodology, AX, LB and VL; visualization, AX; writing— original draft, AX; writing—review and editing, AX, EE, SG, PB, LB, PABP, VTEA, MCH, KR, MGT and FS. The authors read and approved the final manuscript.

## Acknowledgements

We dedicate this paper in memorial of our research adviser, mentor, and friend, Viviane Labrie. We thank the Van Andel Institute Genomics and Bioinformatics and Biostatistics Cores.

## Authors’ information

**Center for Neurodegenerative Science, Van Andel Institute, Grand Rapids, MI, 49503 USA**

Aoji Xie, Elizabeth Ensink, Peipei Li, Juozas Gordevičius, Lee L. Marshall, Sonia George, Patrik Brundin, Lena Brundin and Viviane Labrie

**Center for Epigenetics, Van Andel Institute, Grand Rapids, MI, 49503 USA**

J. Andrew Pospisilik

**Department of Neurology, Helsinki University Hospital, and Clinicum, University of Helsinki, Haartmaninkatu 4, 00290 Helsinki, Finland**

Velma T. E. Aho and Pedro A. B. Pereira and Filip Scheperjans

**Institute of Biotechnology, DNA Sequencing and Genomics Laboratory, University of Helsinki, Viikinkaari 5D, 00014 Helsinki, Finland**

Velma T. E. Aho, Pedro A. B. Pereira, Lars Paulin, Petri Auvinen

**Nell Hodgson Woodruff School of Nursing, Emory University, 1520 Clifton Rd, Atlanta, GA 30322, USA**.

Madelyn C. Houser

**Department of Physiology, Emory University School of Medicine, 615 Michael St, Atlanta, GA 30322, USA**.

Madelyn C. Houser and Malú G. Tansey

**Faculty of Chemistry, Biotechnology and Food Science (KBM), Norwegian University of Life Sciences, 1433 Oslo, Ås, Norway**

Knut Rudi

**Department of Neuroscience and Neurology, Center for Translational Research in Neurodegenerative Disease, University of Florida College of Medicine, 1149 Newell Dr**., **Gainesville, FL 32611, USA**.

Malú G. Tansey

**Division of Psychiatry and Behavioral Medicine, College of Human Medicine, Michigan State University, Grand Rapids, MI, 49503 USA**

Patrik Brundin, Lena Brundin and Viviane Labrie

## Supplementary Files

Supplementary Figures

