## Supplemental Figures for "Butyrate and related epigenetic changes link Parkinson’s disease to inflammatory bowel disease and depressive symptoms"

a

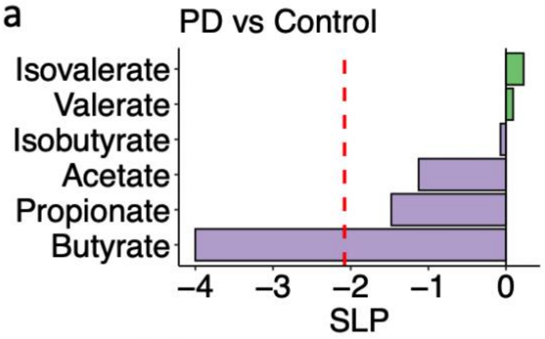


b

| Genus | DiagnosisP | P.value | FDR |
|---|---|---|---|
| Bifidobacterium | 2.19424579 | 1.86E-05 | 0.00019093 |
| Butyricicoccus | -0.733936 | 0.0077645 | 0.03388144 |
| Clostridium_XlVa | -0.7702649 | 0.00704787 | 0.03258728 |
| Lactobacillus | 1.64483732 | 0.0028955 | 0.0160366 |
| Prevotella | -2.1439832 | 0.01055874 | 0.04471937 |
| Roseburia | -1.5899454 | 0.00085493 | 0.00647945 |

c

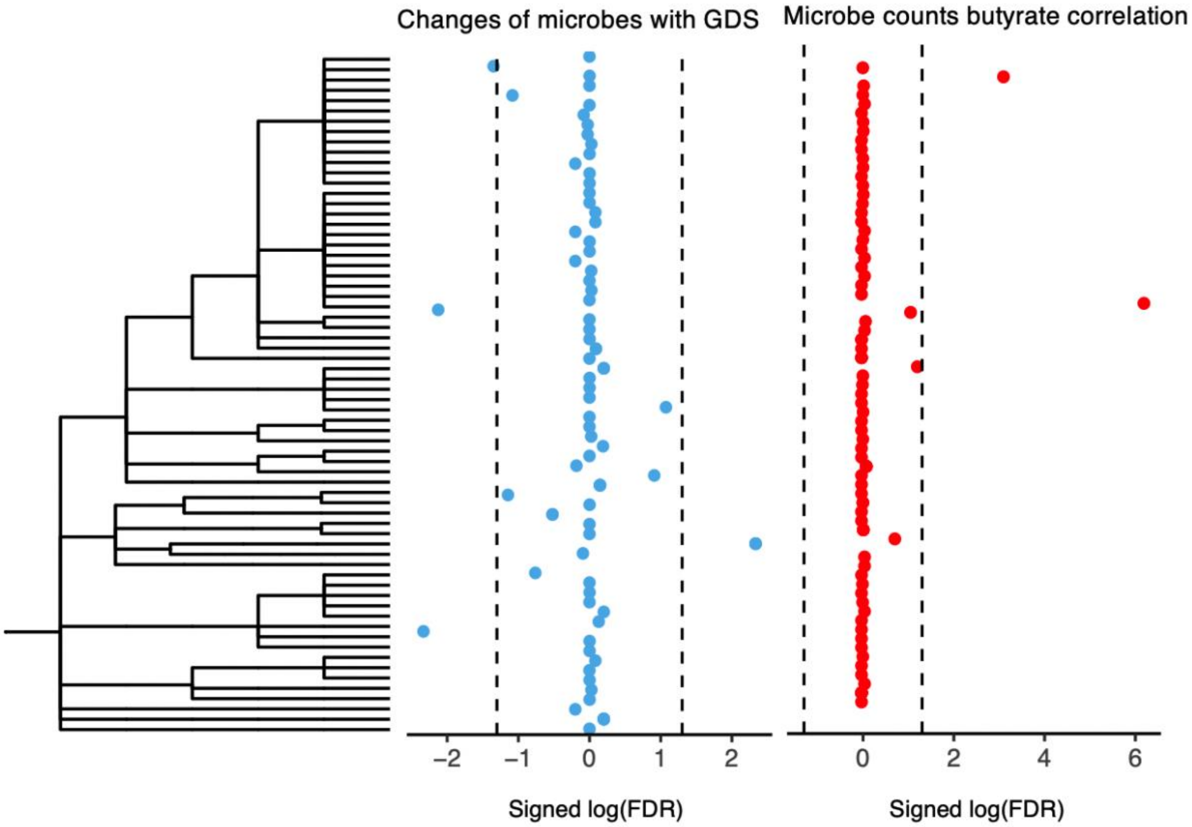


Significant altered microbes with GDS

| Genus | cor GDS | P.value | FDR |
|---|---|---|---|
| Deltaproteobacteria _unclassified | 0.35460432 | 6.28E-05 | 0.00464483 |
| Prevotella | -0.3021576 | 4.37E-05 | 0.00464483 |
| Romboutsia | -0.2881489 | 0.00020207 | 0.00747643 |
| Roseburia | -0.2311234 | 0.00151744 | 0.04491624 |

Microbes significantly correlated with butyrate

| Genus | P.value | Estimate | FDR |
|---|---|---|---|
| Romboutsia | 4.87E-09 | 0.69204318 | 6.47E-07 |
| Roseburia | 1.81E-05 | 0.54335541 | 0.00080422 |

Fig. S1:

a: Short chain fatty acids changes in PD patients relative to controls were identified (robust linear regression, adjusting for age, sex, smoking status and BMI). b: Bacterial genera which were altered in the stool from PD patients relative to control group (Benjamini-Hochberg FDR $q < 0.05$, metagenomeSeq ZIG model). c: Left figure: phylogeny of the 71 bacterial genera included in the analysis. Middle figure: correlations between microbes and GDS (adjusting for age, sex, smoking status and BMI, metagenomeSeq ZIG model). Right figure: correlation between butyrate and genera (read count) in PD patients. Left table: bacteria which significantly correlated with GDS (Benjamini-Hochberg FDR $q < 0.05$, metagenomeSeq ZIG model). Right table: bacteria which significantly correlated with butyrate levels (Benjamini-Hochberg FDR $q < 0.05$, Pearson correlation).

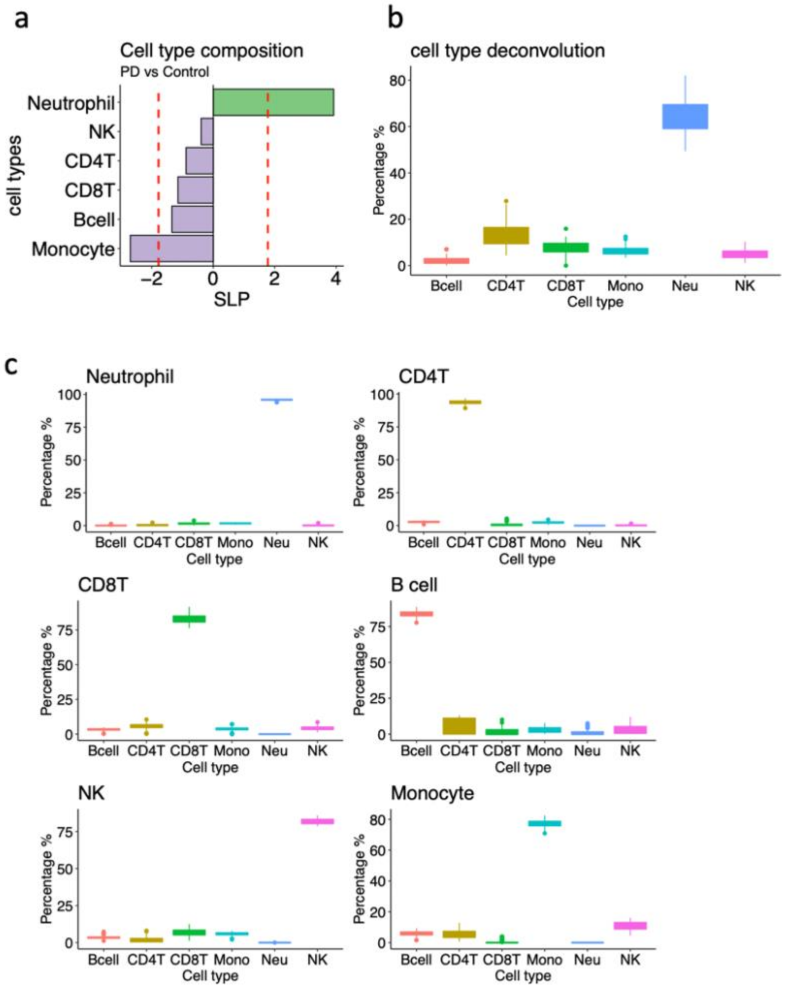


Fig S2:

a: Blood cell percentage in PD compared to control. CIBERSORT was used to perform the cell type decomposition from EPIC methylation array. b: Epigenome of different blood cell types was estimated by Tensor Composition Analysis (TCA) based on the M value of methylated cytosines and the cell percentage. c: For the sanity check, we deconvoluted the estimated epigenome using CIBERSORT again. The corresponding cell type dominates in each epigenome, respectively.

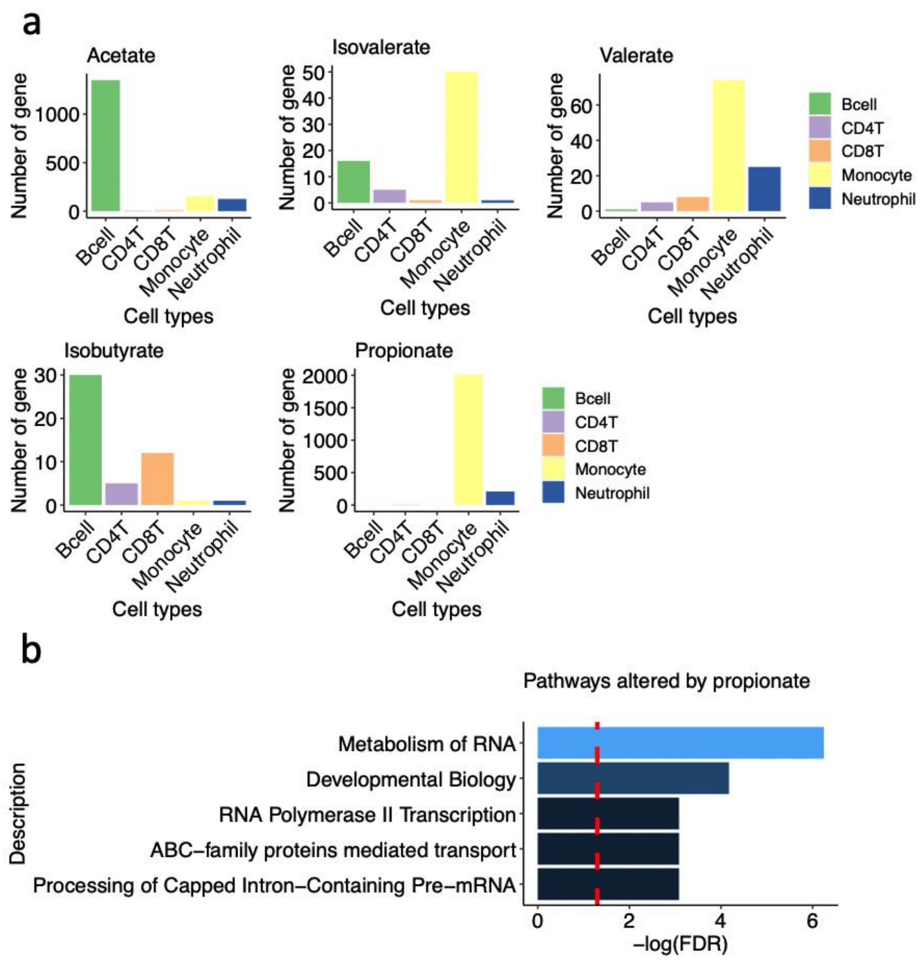


Fig S3:

a: Significant methylation sites (related gene numbers) altered by SCFAs in the epigenome of neutrophils, monocytes, CD8$^+$T cells, CD4$^+$T cells and B cells. b: Gene set enrichment analysis on the neutrophil genes that are epigenetically altered by propionate. Significant threshold Benjamini-Hochberg FDR $q < 0.05$. We found no significant pathways for genes that were epigenetically altered by acetate, isobutyrate, isovalerate and valerate.

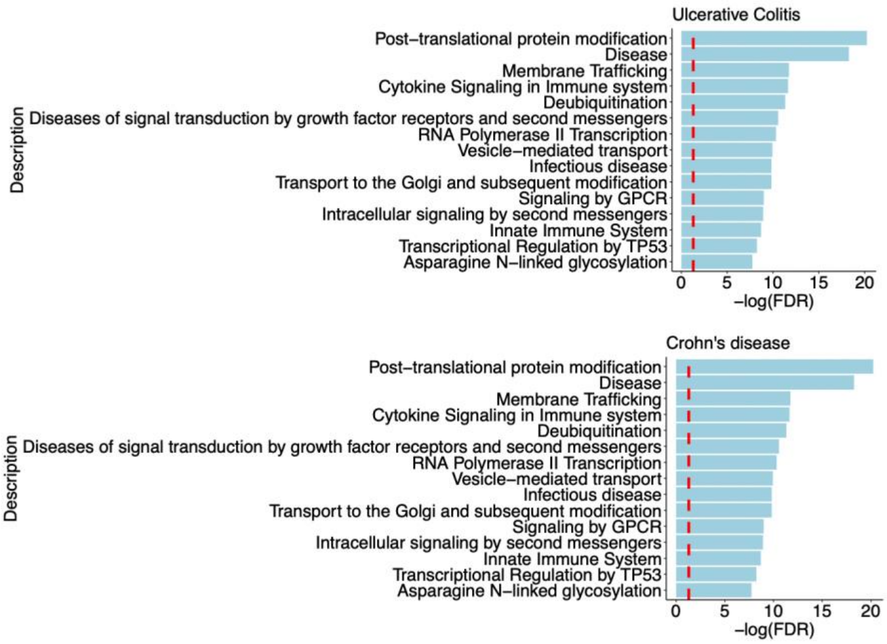


Fig S4: Gene set enrichment analysis on the location of common genetic regions in both Parkinson’s disease, butyrate-associated mDNA regions and ulcerative colitis and Crohn’s disease, respectively. The top 15 significant pathways are shown. Significant threshold Benjamini-Hochberg FDR $q < 0.05$.
